# Teprotumumab Effects on Thyroid Eye Disease in a Prospective Japanese Cohort: MRI-Based Comparison with Intravenous Glucocorticoid Therapy

**DOI:** 10.64898/2026.07.07.26357453

**Authors:** Ichiro Yamauchi, Daisuke Taura, Yohei Ueda, Taku Sugawa, Manabu Miyata, Akinari Yamamoto, Kenji Suda, Eri Nakano, Yo Kishimoto, Koji Nishimura, Yoshitaka Kawai, Mie Abiko, Aya Sakurai, Sadahito Kimura, Daisuke Kosugi, Kentaro Okamoto, Takuro Hakata, Daisuke Yabe

**Affiliations:** Department of Diabetes, Endocrinology and Nutrition, Kyoto University Graduate School of Medicine; Department of Ophthalmology and Visual Sciences, Kyoto University Graduate School of Medicine; Department of Otolaryngology-Head and Neck Surgery, Kyoto University Graduate School of Medicine

**Author notes:** corresponding author: **Address correspondence to:** Ichiro Yamauchi, Department of Diabetes, Endocrinology and Nutrition, Kyoto University Graduate School of Medicine, 54 Kawaharacho, Shogoin, Sakyo-ku, Kyoto 606-8507, Japan.

**Keywords:** Thyroid eye disease, Teprotumumab, Glucocorticoid, Magnetic resonance imaging, Extraocular muscle

## Abstract

**Context:** Teprotumumab (TEP) is an emerging treatment for thyroid eye disease (TED), but real-world evidence outside the United States remains limited, and detailed changes in orbital components have not been fully clarified.

**Objective:** To evaluate the effectiveness of TEP based on clinical manifestations and magnetic resonance imaging (MRI) findings, and compare it with that of intravenous glucocorticoid (IVGC) therapy.

**Methods:** The TEP cohort included all 18 patients who started TEP therapy at Kyoto University Hospital by July 31, 2025. A historical IVGC cohort included 20 patients matched to the TEP cohort.

**Results:** During 24 weeks of TEP therapy, proptosis measured using a Hertel exophthalmometer improved from 22 (20–22) to 19 (16–21) mm (p = 0.025), and clinical activity score decreased from 4 (3–5) to 1 (0–1) point (p < 0.001). Among 15 patients with diplopia, a reduction of at least 1 point in Gorman score was observed in 9 patients (60.0%). Thyroid-stimulating antibody titers decreased from 1,180% (349–4,710) to 282% (132–504) (p = 0.013). MRI-based comparisons with the IVGC cohort showed that TEP reduced both extraocular muscle and orbital fat areas, whereas IVGC reduced extraocular muscle area but conversely increased orbital fat area. Inflamed extraocular muscles identified on MRI were enlarged at baseline and showed marked shrinkage after both therapies.

**Conclusion:** TEP showed robust effectiveness in Japanese real-world patients with TED. MRI-based analyses revealed distinct effects of TEP and IVGC on orbital fat and identified inflamed extraocular muscles as treatment-responsive components.

## Introduction

Thyroid eye disease (TED) is a disorder characterized by ocular symptoms and abnormalities that most commonly occur in associations with Graves’ disease. The pathophysiology of TED involves inflammation and remodeling of orbital tissues, which develop through the accumulation of glycosaminoglycans, adipogenesis, and immune activation mediated by cytokines and chemokines (1). These processes, driven mainly by orbital fibroblasts, cause edema, extraocular muscle enlargement, and orbital fat expansion, leading to clinical manifestations such as proptosis, ocular pain, eyelid swelling and retraction, restricted eye movement, and diplopia. Thus, TED alters facial appearance, disturbs visual function, and impairs quality of life.

For patients with moderate-to-severe TED, systemic immunosuppressive therapy is generally used to control active inflammation (2–4). Intravenous glucocorticoid (IVGC) therapy has long been a standard treatment, with evidence supporting its superiority over oral glucocorticoids (5). Orbital radiotherapy, local glucocorticoid injection, and immunosuppressive agents are also used in selected cases. Once active inflammation has subsided, rehabilitative surgery may be considered to improve residual ocular dysfunction and altered facial appearance.

However, IVGC therapy has several limitations. Although glucocorticoids are effective in improving pain and periocular edema, they do not sufficiently improve proptosis or diplopia in a substantial number of patients. Recently, teprotumumab (TEP), a fully human monoclonal antibody that inhibits IGF-1 receptor, has been developed as a therapeutic agent for TED (6). TSH receptor is a well-established autoantigen involved in TED, as reflected by the association between TED activity and TSH receptor antibody titers (7,8). IGF-1 receptor is co-expressed with TSH receptor in orbital fibroblasts, and signaling crosstalk between these receptors has been demonstrated (9,10). Based on clinical trial evidence, TEP was approved for TED in the United States in 2020 and in Japan in 2024. In clinical trials, TEP significantly improved proptosis, clinical activity score (CAS), and diplopia (11–13). The American Thyroid Association and European Thyroid Association consensus statement recommends TEP as one of the preferred therapies for TED (3,4).

Although the use of TEP is expected to expand to additional countries and regions, several clinical questions remain unresolved. In particular, real-world evidence outside the United States is limited. TED manifestations vary across ethnic groups, and differences in orbital anatomy result in milder proptosis in Asian populations (14,15). Meanwhile, it has not been fully elucidated how TEP affects individual orbital components, such as extraocular muscles and orbital fat. Furthermore, comparisons of the therapeutic effects of TEP with those of IVGC are extremely limited, and few studies have not objectively evaluated orbital components using imaging modalities such as computed tomography (CT) or magnetic resonance imaging (MRI) (16,17).

We launched a prospective observational study of patients with TED, with a particular focus on TEP therapy. A major strength of our study is MRI-based assessment that enabled us to assess changes in extraocular muscles and other orbital components separately and to visualize their inflammation. In the present study, we investigated the clinical effectiveness of TEP in real-world practice in Japan and compared its effects on orbital components with those of IVGC using an institutional historical cohort.

## Materials and Methods

### Patients

The TEP cohort consisted of all 18 patients who started TEP therapy at Kyoto University Hospital between January 1, 2025 and July 31, 2025. All patients completed 24 weeks of follow-up, including those who discontinued TEP therapy. This prospective observational study was approved by the Institutional Review Board and Ethics Committee of the Kyoto University Graduate School of Medicine (approval number: R4795), and written informed consent was obtained from all participants.

For comparison, we retrospectively identified patients who received IVGC therapy at Kyoto University Hospital. Instead of obtaining written informed consent, we provided each patient with the opportunity to opt out of the study through our website. This retrospective analysis was approved by the Institutional Review Board and Ethics Committee of the Kyoto University Graduate School of Medicine (approval number: R3075). The study was conducted in accordance with the principles of the Declaration of Helsinki.

### TEP administration and clinical assessments

TEP was administered intravenously according to the approved dosage and interval. Briefly, we administered TEP at 10 mg/kg over 90 min for the first infusion, 20 mg/kg over 90 min for the second infusion, and 20 mg/kg over 60 min from the third infusion to the final infusion.

Blood tests were performed before TEP administration and at 3, 6, 12, and 24 weeks thereafter, designated as weeks 0, 3, 6, 12, and 24, respectively. Serum levels of TSH and free T4 (fT4) were measured using the Elecsys kits (Roche Diagnostics, Mannheim, Germany; the reference ranges were 0.500–5.000 μIU/mL and 0.880–1.620 ng/dL, respectively). TSH receptor antibody (TRAb) was measured using the AIA kit (Tosoh, Tokyo, Japan), and thyroid-stimulating antibody (TSAb) was measured using the Yamasa Biosensor kit (Yamasa, Chiba, Japan): reference ranges were < 2.0 IU/L and < 110%, respectively. Clinical activity score (CAS) was initially determined by physicians and subsequently reviewed by other physicians based on patients’ symptoms and photographs. Audiometry was routinely performed at weeks 0, 12, and 24, and additionally when patients reported hearing-related symptoms.

### MRI assessments

MRI was performed before TEP administration and at weeks 12 and 24. Proptosis was assessed using a Hertel exophthalmometer in routine clinical practice; however, for the present study, proptosis was further assessed using axial MRI images to ensure objective comparisons (Figure 1A). The dominant eye was defined as the eye with more severe proptosis. To assess longitudinal changes in extraocular muscles and orbital fat, we analyzed T1-weighted coronal images with a slice thickness of 0.9 mm. Images obtained three slices posterior to the posterior pole of the eye were used for analysis. Total orbital area, extraocular muscle area, and optic nerve area were measured separately, and orbital fat area was calculated by subtracting extraocular muscle area and optic nerve area from total orbital area (Figure 1B, 1C). Extraocular muscle inflammation was determined based on increased signal intensity on T2 Dixon water-phase images or T2 short tau inversion recovery (STIR) images. T2 Dixon water-phase images were used in 16 patients, and STIR images were used in 2 patients. A representative image of extraocular muscle inflammation is shown in Figure 1D.

**Figure 1.**
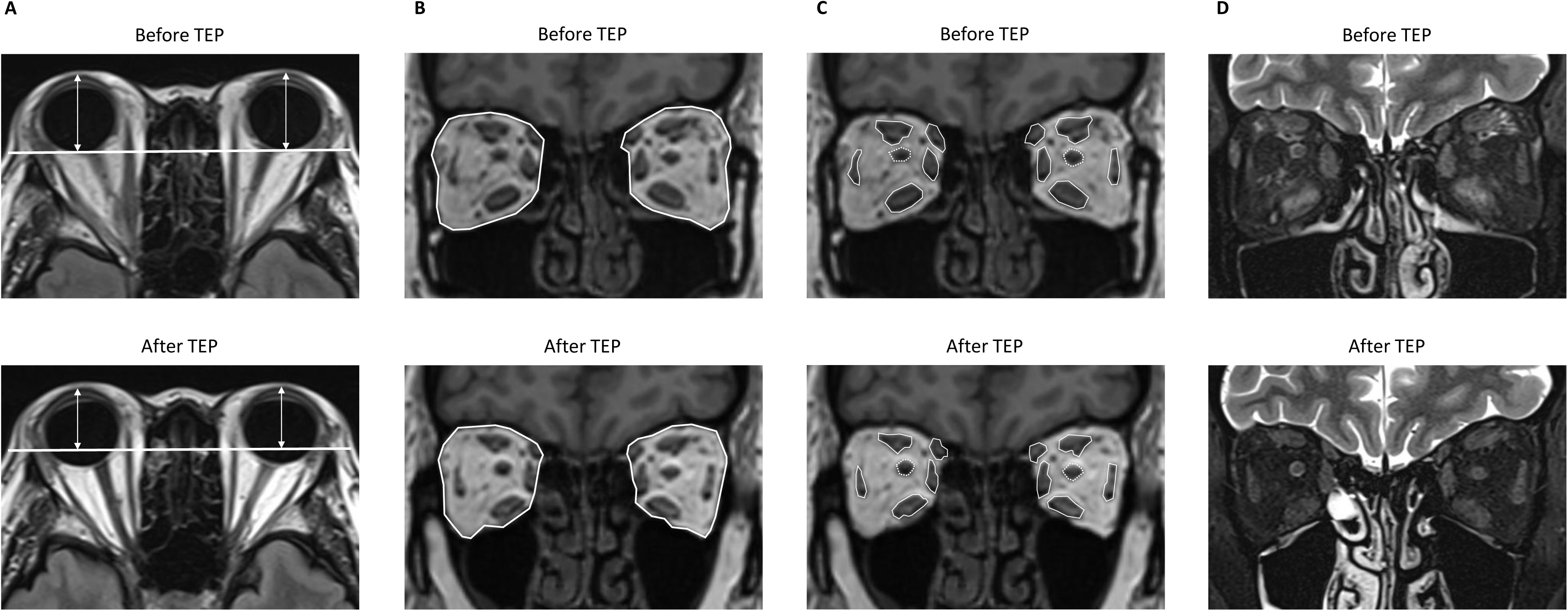
MRI-based assessment in the present study. (A) Measurement of proptosis on T2 FLAIR axial images. (B) Measurement of total orbital area on T1-weighted coronal images. (C) Measurement of extraocular muscle area (solid line) and optic nerve area (dotted line). Orbital fat area was calculated by subtracting extraocular muscle and optic nerve areas from total orbital area. (D) Representative image showing inflammation in extraocular muscles as increased signal intensity on T2 Dixon water-phase images. MRI, magnetic resonance imaging; TED, thyroid eye disease; TEP, teprotumumab.

### Historical IVGC cohort

IVGC therapy was principally performed using a daily regimen consisting of methylprednisolone 0.5 g daily for 3 consecutive days during weeks 1–3, followed by oral prednisolone (PSL) (18). Oral PSL was initiated at 20–30 mg/day according to body weight and gradually tapered. All patients concomitantly received orbital radiotherapy at a cumulative dose of 20 Gy in 10 fractions. MRI was initially performed immediately after the final administration of methylprednisolone and was repeated during follow-up.

A total of 54 patients received initial IVGC therapy between January 1, 2019, and July 31, 2025. We excluded patients who subsequently received TEP therapy (n = 7) and those who did not undergo follow-up MRI within 1 year after IVGC therapy (n = 16). To match baseline characteristics with those of the TEP cohort, we additionally excluded current smokers (n = 7) and patients with low CAS, defined as CAS 0 or 1 (n = 4). The remaining 20 patients were included in the historical IVGC cohort. In this cohort, TSAb was measured using the Yamasa Biosensor kit in 5 patients and the Yamasa EIA kit in 15 patients (Yamasa; reference range, < 120%). STIR images were used in 19 patients, and T2 Dixon water-phase images were used in 1 patient to determine extraocular muscle inflammation.

### Statistical analysis

The sample size was determined by the number of patients treated during the study period. Continuous variables are expressed as medians and interquartile ranges. Comparisons of baseline characteristics were performed using the Mann–Whitney U test for continuous variables and the Fisher’s exact test for categorical variables. For longitudinal data, the Steel test was used to compare values at each time point with those at week 0. Correlations were determined using Spearman’s correlation coefficient. P-values of < 0.05 were considered statistically significant. Graphs were generated using GraphPad Prism 11.0.1 (GraphPad Software, Boston, MA, USA). All statistical analyses were performed using JMP Student Edition 19.0.3 (SAS Institute Inc., Cary, NC, USA).

## Results

### Patient characteristics and therapeutic effects of TEP

Characteristics of 18 patients in the TEP cohort are shown in Table 1. The majority of patients were female, and there were no current smokers. A substantial proportion of patients had a history of IVGC (50.0%) and orbital radiation (38.9%). Thyroid function was well controlled with antithyroid drugs (61.1%) or after total thyroidectomy (22.2%). Proptosis, as assessed using a Hertel exophthalmometer, was 22 (20–22) mm in the dominant eye. The median CAS was 4 points; 2 patients with CAS < 3 points were considered to have active TED on inflammatory findings on MRI. Diplopia was observed in 15 patients (83.3%), and 2 patients had optic neuropathy.

**Table 1.** Baseline patient characteristics in the TEP cohort.

|  | **TEP (n = 18)** |
| --- | --- |
| **Age (year)** | 58 (50–69) |
| **Sex (n)** |  |
| Male | 3 (16.7%) |
| Female | 15 (83.3%) |
| **Smoking status (n)** |  |
| Never | 7 (38.9%) |
| Past | 11 (61.1%) |
| Current | 0 (0.0%) |
| **History of TED-related treatment (n)** |  |
| IVGC | 9 (50.0%) |
| Orbital radiation | 7 (38.9%) |
| **Thyroid function and autoantibodies** |  |
| TSH (μIU/mL) | 0.603 (0.035–2.448) |
| fT4 (ng/dL) | 1.115 (0.920–1.450) |
| TRAb (IU/L) | 5.4 (2.0–14.5) |
| TSAb (%) | 1375 (387–4748) |
| **Proptosis of the dominant eye (mm)** | 22 (20–22) |
| **CAS** | 4 (3–5) |
| **Gorman score** | 2 (1–3) |
| 0: Absent (n) | 3 (16.7%) |
| 1: Intermittent (n) | 3 (16.7%) |
| 2: Inconstant (n) | 7 (38.9%) |
| 3: Constant (n) | 5 (27.8%) |

Values are presented as median (interquartile range) or n (%). Proptosis was measured using Hertel exophthalmometer. TEP, teprotumumab; TED, thyroid eye disease; IVGC, intravenous glucocorticoid; fT4, free T4; TRAb, TSH receptor antibody; TSAb, thyroid-stimulating antibody; and CAS, clinical activity score.

TEP therapy, consisting of 8 infusions over 24 weeks, was completed in 12 patients, whereas 6 patients discontinued treatment due to hearing loss: 1 patient discontinued at week 6, and 5 patients discontinued at week 12. Incomplete treatment courses are not uncommon in real-world practice; in a large multicenter study, 101 of 131 patients (77.1%) completed all 8 infusions (19). Diabetes mellitus newly developed in 3 patients without a prior history of diabetes mellitus and worsened in 1 of 3 patients who had been receiving glucose-lowering agents. However, no patients discontinued TEP therapy due to diabetes mellitus.

We observed significant effects of TEP on symptoms and clinical manifestations. Proptosis improved from 22 (20–22) mm at week 0 to 19 (17–21) mm at week 12 (p = 0.018) and 19 (16–21) mm at week 24 (p = 0.025) (Figure 2A). CAS decreased from 4 (3–5) points at week 0 to 1 (0–1) point at week 12 (p < 0.001) and week 24 (p < 0.001) (Figure 2B). The number of patients achieving CAS of 0 or 1 point reached 14 (77.8%) by week 12 and remained unchanged at week 24. Diplopia also improved during TEP therapy: among the 15 patients with diplopia at baseline, a reduction of at least 1 point in the Gorman score was observed in 9 patients (60.0%), and complete resolution of diplopia was achieved in 3 patients (20.0%) (Figure 2C).

**Figure 2.**
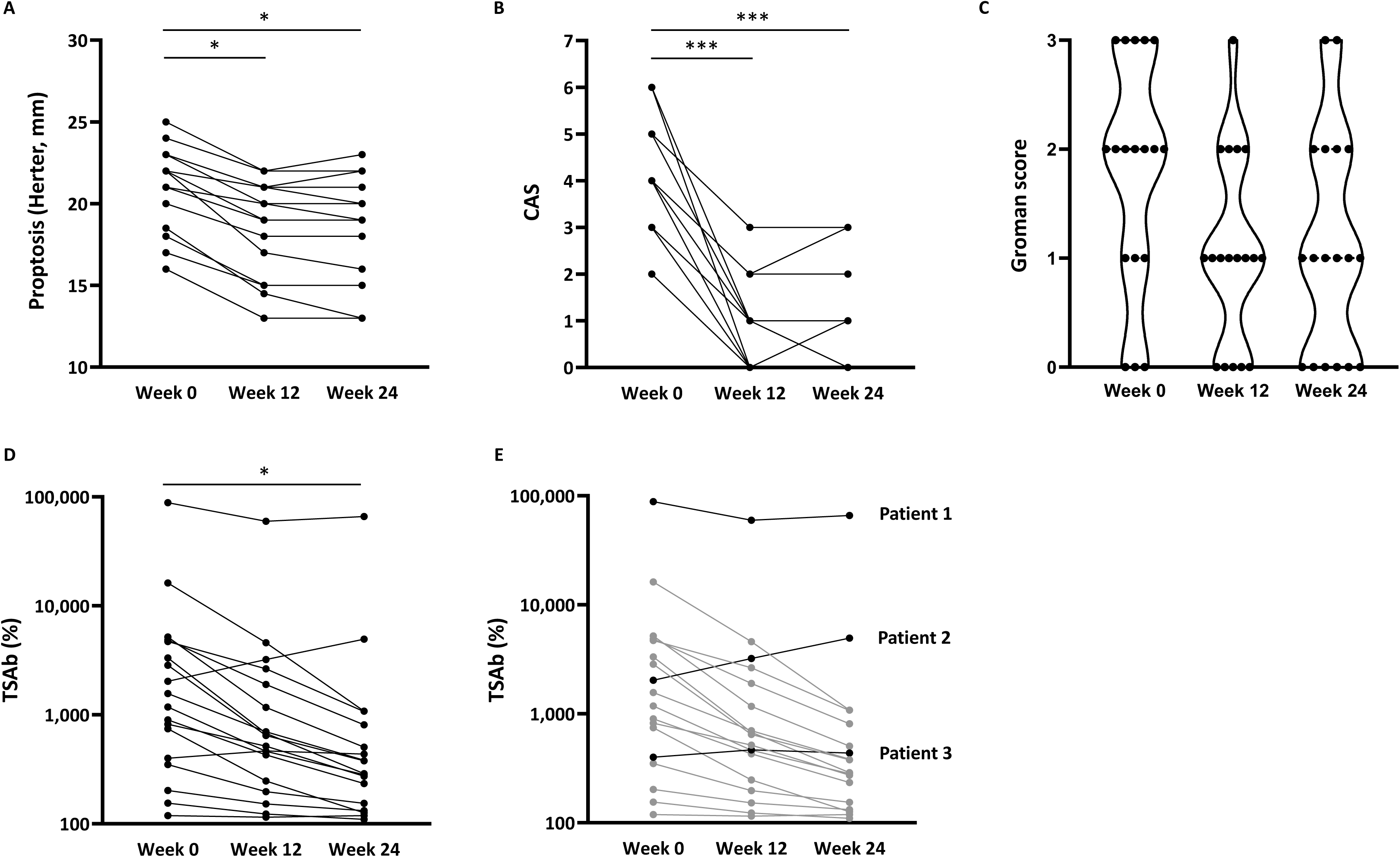
Clinical courses of TED in the TEP cohort. (A) Changes in proptosis measured using a Hertel exophthalmometer. (B) Changes in clinical activity score (CAS). (C) Changes in Gorman score. (D, E) Changes in thyroid-stimulating antibody (TSAb) titers. Panel E highlights 3 patients with mild or no decrease in TSAb titers. Statistical analyses were performed using the Steel test, with week 0 as the reference. *p < 0.05, ***p < 0.001.

TSAb titers gradually decreased from 1,180% (349–4,710) at week 0 to 516% (197–1,170) at week 12 (p = 0.111) and 282% (132–504) at week 24 (p = 0.013) (Figure 2D). However, TSAb titers showed only mild or no decrease in 3 patients, as indicated in Figure 2E. Patient 1 had an extremely high TSAb titer of 88,400%, and her TRAb titer exceeded 40.0 IU/L. Furthermore, TSH receptor-blocking antibody activity was detected using the TSBAb Yamasa kit, with a value of 98.5% (reference range, < 40.0%). Patient 2 exhibited a discrepancy between TSAb and TRAb results: TSAb was 2,030%, whereas TRAb was < 0.9 IU/L. Patient 3 discontinued TEP therapy at week 6 due to hearing loss. Although TSAb changes in these patients should be interpreted cautiously because of blocking activity, assay-related issues, and differences in treatment duration, TSAb titers generally decreased during TEP therapy.

### Characterization of the historical IVGC cohort

We subsequently compared the therapeutic effects of TEP with those of IVGC. According to the criteria described in Materials and Methods, the baseline characteristics of the historical IVGC cohort were matched to those of the TEP cohort, including CAS and Gorman score (Table 2). In the IVGC cohort, MRI was performed before and immediately after IVGC and during the follow-up period within 1 year. Follow-up MRI was performed at 176 (167–184) days in the TEP cohort and 200 (152–259) days in the IVGC cohort, with no substantial difference between the cohorts.

**Table 2.**
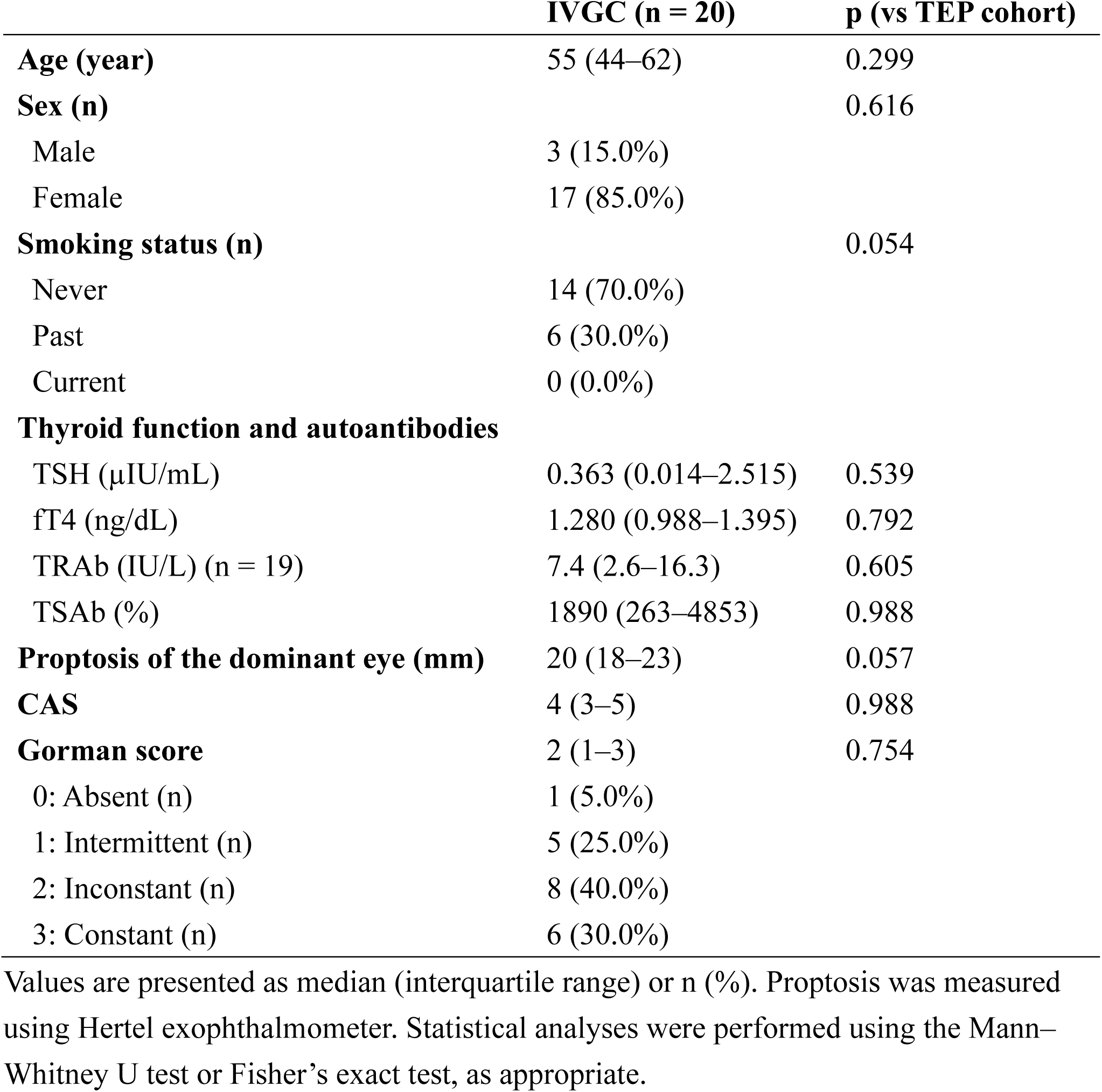
Baseline patient characteristics in the historical IVGC cohort.

IVGC provided clinical effects on TED similar to those of TEP, except for proptosis. CAS exhibited comparable decreases in both cohorts: 1 (0–1) point at week 24 in the TEP cohort and 1 (0–2) point during the follow-up period in the IVGC cohort (p = 0.159). Among patients with diplopia at baseline, a reduction of at least 1 point in the Gorman score was observed in 8 of 18 patients (44.4%) in the IVGC cohort, with no significant difference compared with the TEP cohort (p = 0.295). TSAb titers also decreased after IVGC: the percent change in TSAb was −74.8% (−84.2 to −28.1) at week 24 in the TEP cohort and −46.8% (−90.9 to −1.5) during follow-up in the IVGC cohort (n = 18; p = 0.800). In the IVGC cohort, oral PSL was continued during the follow-up period in 11 patients (55.0%): 6 patients received 10 mg/day, 1 patient received 7.5 mg/day, and 4 patients received 5 mg/day.

### Changes in orbital fat and extraocular muscles during TEP and IVGC therapies

In contrast,to changes in proptosis differed significantly between the two cohorts. Proptosis measured on MRI showed sustained improvement in the TEP cohort. In the IVGC cohort, proptosis improved immediately after IVGC but partially worsened during the follow-up period (Figure 3A, 3B, Table 3). Consequently, the final change in proptosis differed significantly between the TEP and IVGC cohorts (Figure 3C): −3.4 mm (−4.6 to −2.5) and −0.7 mm (−1.5 to 0.0), respectively (p < 0.001). No patients in the IVGC cohort showed an improvement of 2 mm or more, whereas 16 of 18 patients (88.9%) in the TEP cohort did.

**Figure 3.**
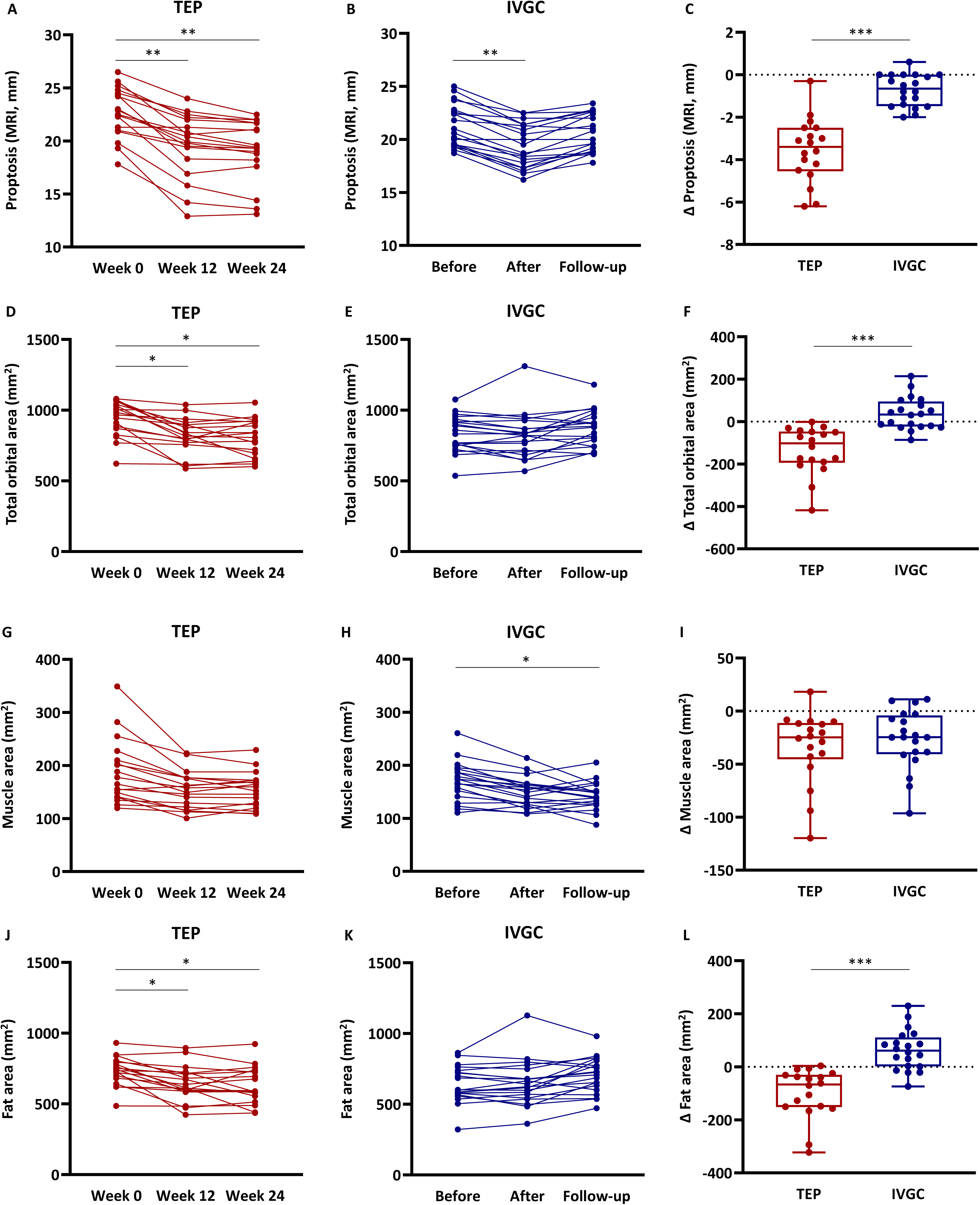
MRI-based comparisons between TEP and IVGC therapies. (A–C) MRI-based proptosis in the TEP cohort (A) and IVGC cohort (B), and comparison of the final change (Δ) between the cohorts (C). (D–F) Total orbital area in the TEP cohort (D) and IVGC cohort (E), and comparison of the final change (Δ) between the cohorts (F). (G–I) Extraocular muscle area in the TEP cohort (G) and IVGC cohort (H), and comparison of the final change (Δ) between the cohorts (I). (J–L) Orbital fat area in the TEP cohort (J) and IVGC cohort (K), and comparison of the final change (Δ) between the cohorts (L). Final changes were calculated from week 0 to week 24 in the TEP cohort and from before treatment to follow-up in the IVGC cohort. Statistical analyses were performed using the Steel test, with week 0 as the reference, for longitudinal changes and using the Mann–Whitney U test for comparisons of final changes between the cohorts. *p < 0.05, **p < 0.01, ***p < 0.001.

**Table 3.** Summary of MRI-based parameters.

|  | TEP (n = 18) | IVGC (n = 20) | p |
| --- | --- | --- | --- |
| **Week 0 (TEP), Before IVGC (IVGC)** |  |  |  |
| Proptosis (mm) | 22.7 (21.0–24.6) | 20.8 (19.5–22.8) | 0.057 |
| Total orbital area (mm<sup>2</sup>) | 975.7 (859.6–1046.6) | 845.5 (749.8–930.9) | **0.011** |
| Muscle area (mm<sup>2</sup>) | 171.1 (140.6–214.1) | 170.1 (143.9–191.3) | 0.569 |
| Inferior rectus | 40.3 (32.3–62.6) | 52.9 (35.8–64.6) | 0.342 |
| Superior rectus | 40.7 (31.3–59.7) | 39.2 (25.6–44.5) | 0.225 |
| Medial rectus | 41.7 (33.5–52.2) | 32.4 (27.5–40.9) | 0.072 |
| Lateral rectus | 27.4 (23.8–29.9) | 26.3 (22.4–31.0) | 0.640 |
| Superior oblique | 21.5 (12.9–26.9) | 17.9 (13.0–20.1) | 0.165 |
| Fat area (mm<sup>2</sup>) | 739.7 (672.0–797.4) | 599.9 (570.3–734.7) | **0.024** |
| **Week 12 (TEP), After IVGC (IVGC)** |  |  |  |
| Proptosis (mm) | 20.2 (18.0–22.1) | 19.1 (17.4–21.4) | 0.599 |
| Total orbital area (mm<sup>2</sup>) | 826.0 (767.9–900.8) | 826.8 (709.5–913.9) | 0.872 |
| Muscle area (mm<sup>2</sup>) | 153.6 (120.2–177.1) | 151.4 (127.1–164.5) | 0.770 |
| Inferior rectus | 35.1 (20.3–42.0) | 42.9 (26.5–47.7) | 0.286 |
| Superior rectus | 35.9 (29.5–41.2) | 36.7 (27.5–42.7) | 0.988 |
| Medial rectus | 33.5 (30.0–42.5) | 32.0 (26.3–35.6) | 0.236 |
| Lateral rectus | 25.9 (22.7–31.3) | 24.5 (21.6–28.1) | 0.474 |
| Superior oblique | 18.3 (16.0–23.8) | 15.6 (12.3–18.8) | 0.144 |
| Fat area (mm<sup>2</sup>) | 626.2 (587.9–717.7) | 633.9 (549.2–731.8) | 0.942 |
| **Week 24 (TEP), Follow-up (IVGC)** |  |  |  |
| Proptosis (mm) | 19.4 (18.1–21.7) | 20.4 (18.8–22.6) | 0.165 |
| Total orbital area (mm<sup>2</sup>) | 823.7 (686.4–918.8) | 882.6 (754.7–971.3) | 0.260 |
| Muscle area (mm<sup>2</sup>) | 156.0 (124.5–172.2) | 144.1 (126.5–160.8) | 0.320 |
| Inferior rectus | 35.2 (27.6–44.9) | 38.9 (28.5–50.1) | 0.405 |
| Superior rectus | 35.2 (27.0–39.6) | 28.0 (24.5–37.0) | 0.121 |
| Medial rectus | 35.6 (24.0–40.8) | 32.9 (26.8–37.1) | 0.421 |
| Lateral rectus | 28.1 (25.4–33.8) | 22.8 (20.7–28.4) | **0.006** |
| Superior oblique | 18.1 (15.6–21.5) | 15.6 (11.1–19.9) | 0.121 |
| Fat area (mm<sup>2</sup>) | 591.9 (544.2–731.1) | 717.0 (608.5–800.3) | 0.093 |

Values are presented as median (interquartile range). Data from the dominant eyes were used. Statistical analyses were performed using the Mann–Whitney U test. Bold values indicate statistical significance. MRI, magnetic resonance imaging.

We further assessed changes in orbital components. Total orbital area significantly decreased in the TEP cohort, whereas it did not change in the IVGC cohort (Figure 3D, 3E, Table 3). Accordingly, the final change in total orbital area differed significantly between the TEP cohort and the IVGC cohort (Figure 3F): −102.4 mm^2^ (−193.7 to −46.7) and 33.3 mm^2^ (−23.1 to 95.1), respectively (p < 0.001). Extraocular muscle area tended to decrease in both cohorts, although the decrease was not significant in the TEP cohort (Figure 3G, 3H, Table 3). There was no significant difference in the final change in extraocular muscle area between the TEP cohort and the IVGC cohort (Figure 3I): −24.7 mm^2^ (−45.3 to −11.2) and −24.7 mm^2^ (−40.6 to −4.2), respectively (p = 0.567). Orbital fat area significantly decreased during TEP therapy, whereas it increased during the follow-up period after IVGC therapy (Figure 3J, 3K, Table 3). The final change in orbital fat area differed significantly between the TEP cohort and the IVGC cohort (Figure 3L): −66.3 (−151.8 to −30.0) mm^2^ and 61.3 (1.8 to 110.9) mm^2^, respectively (p < 0.001).

### Clinical significance of inflammatory findings in extraocular muscles

Furthermore, we evaluated the clinical significance of inflammation in the extraocular muscles. As shown in Figure 1D, inflammation in each extraocular muscle was identified using Dixon water-phase or STIR images. Extraocular muscles with inflammation were larger than those without inflammation, except for the lateral rectus muscles, and showed marked shrinkage after both therapies (Figure 4A–4D). We also observed negative correlations between pretreatment muscle area and its final change (Figure 4E–4H). These correlations were stronger in inflamed muscles than in non-inflamed muscles in both the TEP cohort (ρ = −0.926 vs. −0.422, both p < 0.001) and the IVGC cohort (ρ = −0.705 vs. −0.341, both p < 0.001). The correlations were not significant in several non-inflamed muscles, as detailed in the Figure 4 legend.

**Figure 4.**
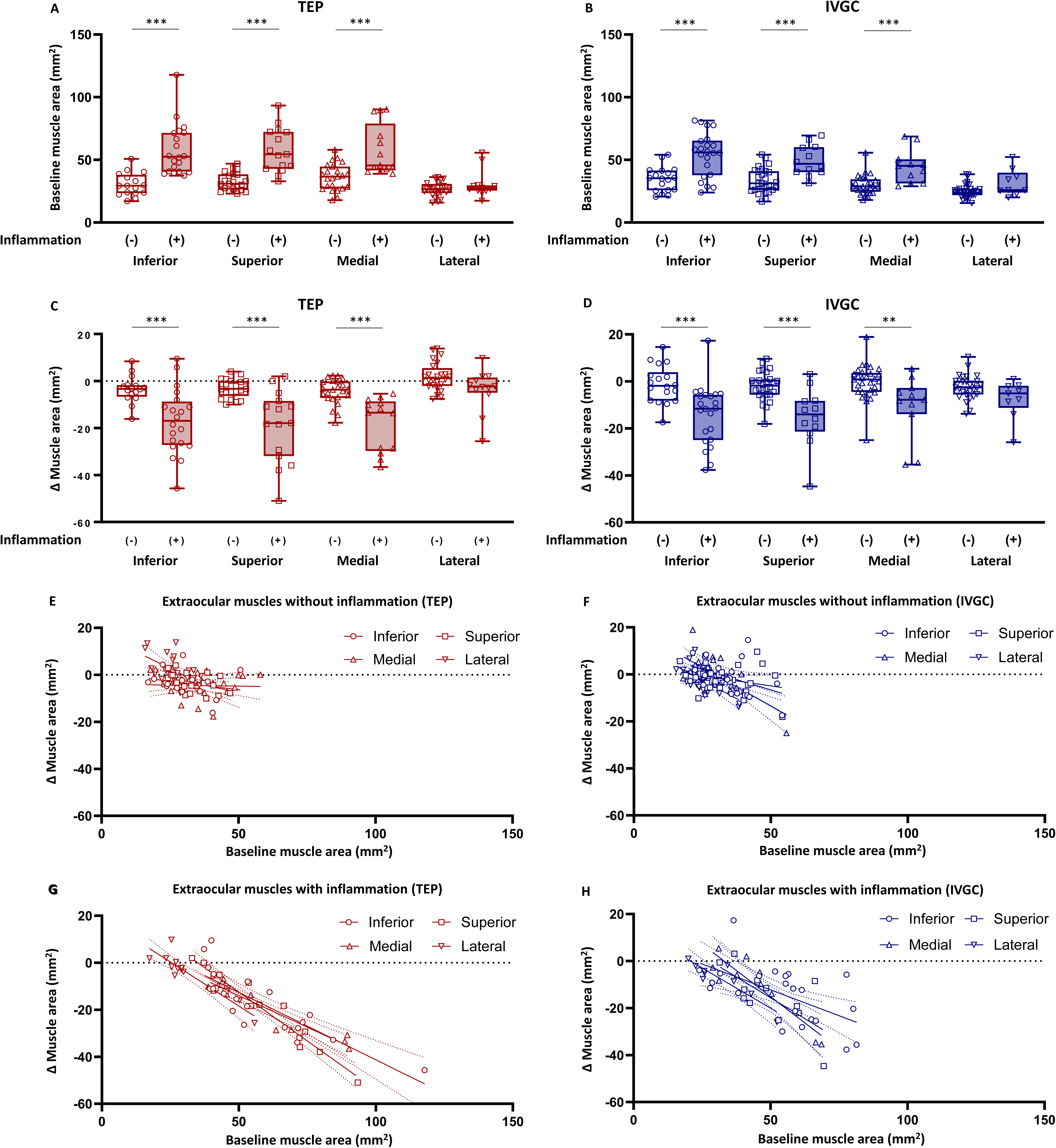
Inflammation-based analyses of extraocular muscles. (A, B) Baseline areas of extraocular muscles with and without inflammation in the TEP cohort (A) and IVGC cohort (B). Extraocular muscles were individually analyzed as the inferior rectus (open circles), superior rectus (open squares), medial rectus (open triangles), and lateral rectus muscles (open inverted triangle). (C, D) Final changes in extraocular muscle area after TEP therapy (C) and IVGC therapy (D) according to the presence or absence of inflammation. (E, F) Correlations between baseline extraocular muscle area and its final change in muscles without inflammation in the TEP cohort (E) and IVGC cohort (F). Correlation coefficients for individual muscles were as follows: in the TEP cohort, ρ = −0.236 (p = 0.380) for the inferior rectus, ρ = −0.413 (p = 0.063) for the superior rectus, ρ = −0.102 (p = 0.644) for the medial rectus, ρ = −0.533 (p = 0.007) for the lateral rectus; in the IVGC cohort, ρ = −0.414 (p = 0.088) for the inferior rectus, ρ = −0.235 (p = 0.228) for the superior rectus, ρ = −0.465 (p = 0.010) for the medial rectus, ρ = −0.620 (p < 0.001) for the lateral rectus. (G, H) Correlations between baseline extraocular muscle area and its final change in muscles with inflammation in the TEP cohort (G) and IVGC cohort (H). Correlation coefficients for individual muscles were as follows: in the TEP cohort, ρ = −0.844 (p < 0.001) for the inferior rectus, ρ = −0.936 (p < 0.001) for the superior rectus, ρ = −0.863 (p < 0.001) for the medial rectus, ρ = −0.832 (p < 0.001) for the lateral rectus; in the IVGC cohort, ρ = −0.635 (p = 0.015) for the inferior rectus, ρ = −0.643 (p = 0.024) for the superior rectus, ρ = −0.818 (p = 0.002) for the medial rectus, ρ = −0.783 (p = 0.013) for the lateral rectus. Inflammation was defined as increased signal intensity on T2 Dixon water-phase or STIR images. Statistical analyses were performed using the Mann–Whitney U test for comparisons between extraocular muscles with and without inflammation and Spearman’s correlation coefficient for correlation analyses. **p < 0.01, ***p < 0.001.

We additionally focused on patterns of extraocular muscle inflammation and analyzed patients with at least one inflamed muscle. MRI images of representative cases with only one inflamed muscle, defined as the isolated type (n = 12; 5 treated with TEP and 7 treated with IVGC), and those with two or more inflamed muscles, defined as the multiple type (n = 19; 9 treated with TEP and 10 treated with IVGC), are shown in Figure 5A. Regarding patient characteristics, the isolated type exhibited milder proptosis, lower TRAb and TSAb titers, and lower CAS than the multiple type, whereas a substantial proportion of patients had diplopia in both types (Table 4).

**Figure 5.**
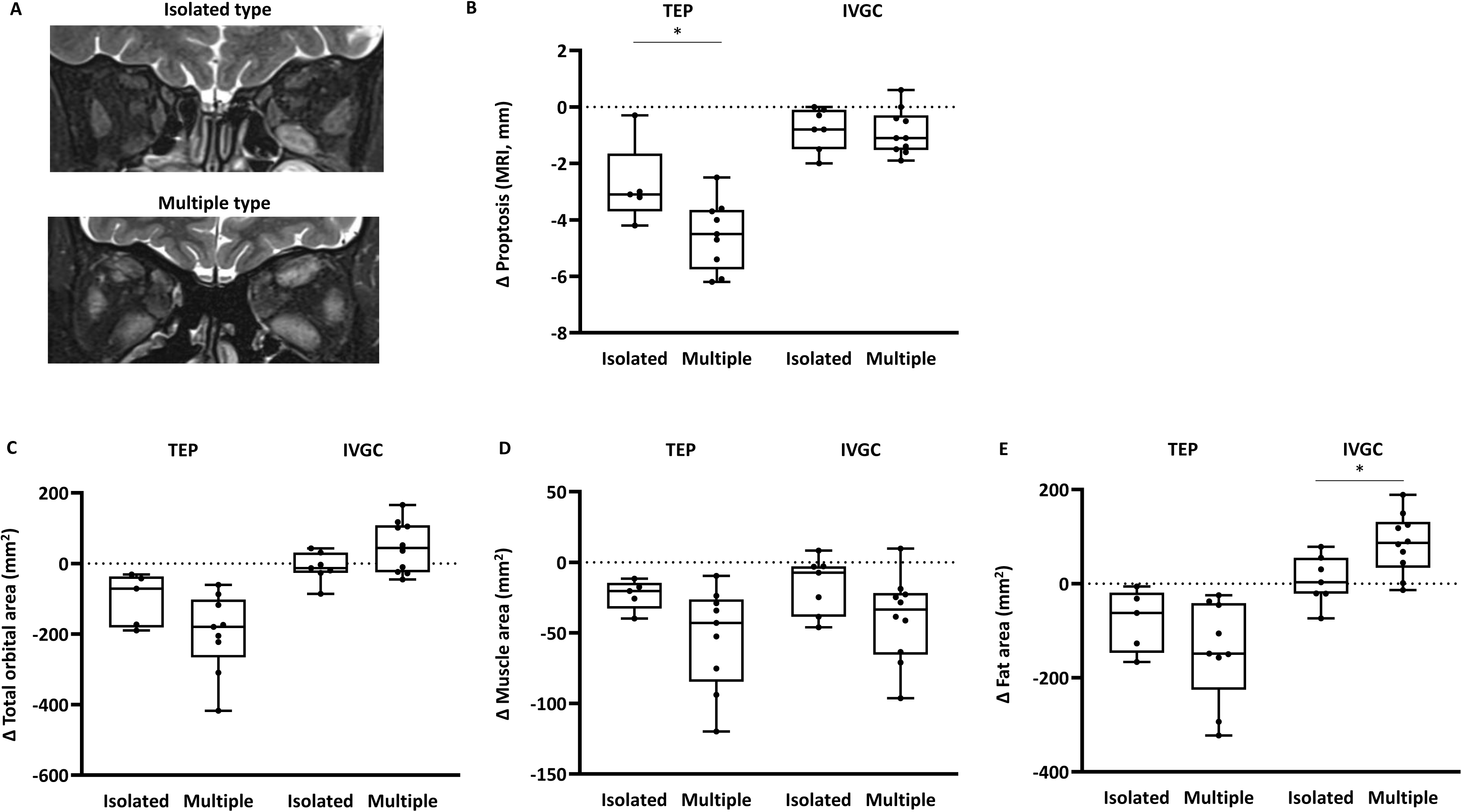
Comparative analyses according to the inflamed muscle pattern. (A) Representative MRI images of the isolated and multiple types. (B) Changes in MRI-based proptosis after TEP or IVGC therapy. (C–E) Final changes in total orbital area (C), extraocular muscle area (D), and orbital fat area (E). The isolated type was defined as involvement of only one inflamed extraocular muscle, and the multiple type as involvement of two or more inflamed extraocular muscles. Statistical analyses were performed using the Mann–Whitney U test. *p < 0.05.

**Table 4.** Comparisons of patient characteristics according to the pattern of extraocular muscle inflammation.

|  | Isolated type (n = 12) | Multiple type (n = 19) | p |
| --- | --- | --- | --- |
| **Treatment (n)** |  |  | 0.552 |
| TEP | 5 (41.7%) | 9 (47.4%) |  |
| IVGC | 7 (58.3%) | 10 (52.6%) |  |
| **Age (year)** | 60 (54–65) | 54 (48–72) | 0.761 |
| **Sex (n)** |  |  | 0.658 |
| Male | 2 (16.7%) | 3 (15.8%) |  |
| Female | 10 (83.3%) | 16 (84.2%) |  |
| **Smoking status (n)** |  |  | 0.061 |
| Never | 4 (33.3%) | 13 (68.4%) |  |
| Past | 8 (66.7%) | 6 (31.6%) |  |
| Current | 0 (0.0%) | 0 (0.0%) |  |
| **Thyroid function and autoantibodies** |  |  |  |
| TSH (μIU/mL) | 0.958 (0.376–4.742) | 0.133 (0.011–2.230) | 0.065 |
| fT4 (ng/dL) | 1.105 (0.940–1.308) | 1.280 (0.984–1.410) | 0.429 |
| TRAb (IU/L) | 2.4 (1.4–9.6) | 9.6 (4.5–17.2) | **0.035** |
| TSAb (%) | 485 (182–1275) | 3130 (819–5190) | **0.005** |
| **CAS** | 3 (2–4) | 4 (3–5) | **0.013** |
| **Gorman score** | 2 (1–3) | 2 (2–3) | 0.680 |
| 0: Absent (n) | 1 (8.3%) | 1 (5.3%) |  |
| 1: Intermittent (n) | 2 (16.7%) | 3 (15.8%) |  |
| 2: Inconstant (n) | 4 (33.3%) | 10 (52.6%) |  |
| 3: Constant (n) | 5 (41.7%) | 5 (26.3%) |  |
| **MRI parameters** |  |  |  |
| Proptosis (mm) | 20.3 (19.4–22.3) | 22.9 (20.2–24.3) | **0.041** |
| Total orbital area (mm<sup>2</sup>) | 847.6 (749.8–958.0) | 944.9 (889.8–1040.8) | **0.027** |
| Muscle area (mm<sup>2</sup>) | 145.0 (131.5–163.1) | 201.1 (178.6–227.4) | **< 0.001** |
| Inferior rectus | 35.4 (28.9–44.9) | 54.4 (38.7–71.4) | **< 0.001** |
| Superior rectus | 27.6 (25.5–31.9) | 45.2 (39.6–59.8) | **< 0.001** |
| Medial rectus | 28.8 (25.4–38.1) | 43.3 (34.8–54.7) | **< 0.001** |
| Lateral rectus | 24.4 (21.9–26.9) | 26.3 (23.2–30.2) | 0.082 |
| Superior oblique | 17.9 (13.3–19.0) | 21.4 (12.3–27.0) | 0.453 |
| Fat area (mm<sup>2</sup>) | 661.7 (582.5–774.8) | 729.1 (685.7–763.1) | 0.362 |

Values are presented as median (interquartile range) or n (%). Statistical analyses were performed using the Mann–Whitney U test or Fisher’s exact test, as appropriate. Bold values indicate statistical significance.

Improvement in proptosis after TEP was more pronounced in the multiple type than in the isolated type (Figure 5B): −4.5 mm (−5.8 to −3.7) and −3.1 mm (−3.7 to −1.7), respectively (p = 0.046). In contrast, improvement in proptosis after IVGC did not differ between the types (Figure 5B): −1.1 mm (−1.5 to −0.3) in the multiple type and −0.8 mm (−1.5 to −0.1) in the isolated type (p = 0.732). Changes in CAS were not significantly different between types. In the isolated type, final CAS of 0 or 1 point was achieved in 5 of 5 patients (100.0%) treated with TEP and 5 of 7 patients (71.4%) treated with IVGC (p = 0.318). In the multiple type, the corresponding numbers were 7 of 9 patients (77.8%) treated with TEP and 4 of 10 patients (40.0%) treated with IVGC (p = 0.115). Improvement in diplopia, defined as a reduction of at least 1 point in the final Gorman score, was observed at similar frequencies between the types. In the isolated type, improvement was observed in 3 of 4 patients (75.0%) treated with TEP and 3 of 7 patients (42.9%) with IVGC (p = 0.349). In the multiple type, improvement was observed in 4 of 9 patients (44.4%) treated with TEP and 3 of 8 patients (37.5%) treated with IVGC (p = 0.581). In the MRI analyses, changes in total orbital area and extraocular muscle area did not differ between the types in either therapy (Figure 5C, 5D). However, orbital fat area tended to increase predominantly in the multiple type after IVGC therapy, whereas this tendency was not observed after TEP therapy (Figure 5E).

## Discussion

In the present study, we report findings from our prospective Japanese cohort of patients with TED who received TEP therapy, including MRI-based assessments. TEP exhibited significant clinical effects, accompanied by decreases in TSAb titers. We constructed a historical IVGC cohort matched to the TEP cohort and performed comparative MRI analyses. Proptosis showed sustained improvement after TEP, whereas improvement observed immediately after IVGC was attenuated during follow-up, which appeared to be attributable to an increase in orbital fat. Extraocular muscles with inflammation identified on MRI were enlarged and responded to both TEP and IVGC therapies. Classification according to the number of inflamed muscles revealed that the isolated type had lower TRAb and TSAb titers and CAS than the multiple type, whereas frequency and severity of diplopia were not different in both types. Proptosis improved more markedly in the multiple type after TEP, but not after IVGC. This difference appeared to be attributable to an increase in orbital fat after IVGC, which was observed predominantly in the multiple type.

Evidence regarding efficacy of TEP in Asian populations remains limited to the Japanese phase 3 clinical trial, OPTIC-J (13). Our real-world cohort demonstrated substantial effectiveness of TEP on proptosis, CAS, and diplopia, broadly consistent with the findings of OPTIC-J. Importantly, our cohort included a substantial proportion of patients with prior IVGC and orbital radiation, suggesting that TEP can be effective even in patients who had received conventional therapies. This observation is consistent with previous reports showing that TEP was effective in patients with refractory TED (20,21).

We also observed a gradual decrease in TSAb titers during TEP therapy. A previous report demonstrated decreases in TRAb and thyroid-stimulating immunoglobulin titers during TEP therapy, although detailed assay information and individual trajectories were not provided (22). In the present study, TSAb titers decreased in a time-dependent manner, even in patients with high baseline titers. This finding may suggest that TEP indirectly modifies autoimmune activity in Graves’ disease and TED. In patients with Graves’ disease, IGF-1 receptor expression was increased on peripheral B cells, and IGF-1 enhanced antibody production from peripheral B cells (23). Flow cytometric analysis of peripheral blood samples from a phase 2 clinical trial of TEP showed that the expression of MHC II, CD80, CD86, and PD-L1 on fibrocytes, as well as IL-17A and IFN-γ expression in CD4L T cells, was reduced in the TEP-treated patients (24). Therefore, the observed TSAb decline may reflect immunological changes associated with suppression of IGF-1 receptor signaling by TEP.

MRI has long been used to evaluate TED activity and orbital tissue involvement. Fat-suppressed T2-weighted sequences, including STIR, can detect inflammatory edema in extraocular muscles and predict therapeutic outcome after IVGC therapy (25–27). In the present study, we mainly used Dixon water-phase images to assess inflammation, an approach that has also been used in previous studies of TED (28–30). Dixon-based imaging has been reported to provide fewer artifacts, shorter acquisition time, and higher self-reported reader confidence than conventional fat-suppressed T2-weighted sequences in the assessment of TED (31).

Previous studies have examined orbital changes in TED using CT or MRI, including an imaging substudy of the phase 3 OPTIC trial of TEP (32) and real-world analyses of TEP-treated patients (33–35). To our knowledge, the present study provides the first MRI-based comparative data between TEP and IVGC. A key difference between TEP and IVGC was observed in orbital fat. Consistent with previous reports (32,34), TEP therapy decreased orbital fat area, whereas IVGC therapy conversely increased orbital fat area during follow-up. This increase in orbital fat may explain weaker and less sustained improvement in proptosis after IVGC. Consistent with this interpretation, Kim *et al.* reported that linsitinib, a small-molecule inhibitor of IGF-1 receptor, reduced adipogenesis in TED orbital fibroblasts (36).

Glucocorticoids are well known to affect adipose tissue metabolism (37) and can alter fat distribution, as observed in Cushing syndrome with excessive glucocorticoid action. In addition to moon face, exophthalmos is a recognized clinical manifestation of Cushing syndrome (38). Furthermore, the natural history of TED, as described by Rundle’s curve (39), may involve expansion of orbital fat over time. Because IVGC primarily suppresses acute inflammation and may not directly inhibit adipogenesis, the observed increase in orbital fat might partly reflect the natural course. Taken together, IVGC may ameliorate inflammatory swelling of extraocular muscles without preventing subsequent or ongoing expansion of orbital fat.

MRI-based assessment is routinely performed in accordance with recommendations of the Committee on Diagnostic Criteria and Treatment Guidelines for Thyroid Eye Disease of the Japan Thyroid Association and the Japan Endocrine Society. Analyses of individual extraocular muscles showed that the lateral rectus muscles were rarely enlarged and showed no significant shrinkage after either TEP or IVGC, consistent with a previous report (35). We further elucidated clinical significance of MRI-detected inflammation in extraocular muscles. Our results showed that MRI-detected inflammation was associated with extraocular muscle enlargement and responses to both TEP and IVGC therapies. Importantly, in non-inflamed muscles, the correlation between baseline muscle area and its final change was weaker than in inflamed muscles. Thus, MRI-detected inflammatory status may help predict responses to TEP and IVGC.

Unilateral or asymmetric TED has long been recognized, and a recent multicenter study from the European Group on Graves’ Orbitopathy demonstrated that asymmetric TED was more severe than symmetric TED (40). To further characterize asymmetric or localized TED, we evaluated the clinical features of the isolated type, a single-muscle involvement pattern often identified using MRI or CT. We found that patients with the isolated type had milder proptosis, lower CAS, and lower TRAb and TSAb titers than those with the multiple type. Nevertheless, diplopia severity was comparable between the groups. These findings suggest that diplopia severity is not determined solely by the extent of proptosis or inflammatory activity, but may also be influenced by the localization and asymmetry of extraocular muscle involvement. Importantly, IVGC therapy increased orbital fat area predominantly in the multiple type. Taken together, TEP and IVGC may have similar effectiveness in the isolated type but distinct effects in the multiple type: TEP improved proptosis without increasing orbital fat.

The present study has several strengths. First, we prospectively collected detailed clinical data including TSAb titers and MRI findings, in TEP-treated patients. Second, MRI allowed objective assessment of proptosis and orbital tissue components, namely extraocular muscles and orbital fat. Third, direct comparison of TEP with IVGC provided insight into the anatomical basis of their different effects on proptosis. Finally, we evaluated the clinical significance of MRI-identified inflammation in extraocular muscles.

This study also has several limitations. First, it was conducted at a single center and included a relatively small number of patients. The reproducibility of our observations should be verified in larger multicenter studies including other ethnic groups. Second, the IVGC cohort was retrospective and historical. In particular, the IVGC cohort included only treatment-naïve patients, whereas the TEP cohort included a substantial number of patients with refractory TED. Despite this potential disadvantage, TEP demonstrated substantial clinical effectiveness in this cohort. In addition, the timing of follow-up MRI in the IVGC cohort was not fully consistent, although the follow-up interval did not differ significantly from that in the TEP cohort. Third, TSAb assays changed during the period in which the IVGC cohort was treated, limiting direct comparison of TSAb trajectories between the TEP and IVGC cohorts. Fourth, quantitative assessment of inflammation could not be performed because STIR imaging protocols in the IVGC era were not fully standardized. Dixon-based assessment may help overcome this limitation in future studies. Finally, MRI measurements were based on selected images, and validation using volumetric methods is required.

In conclusion, TEP demonstrated robust effectiveness for TED in Japanese real-world settings, even though our prospective cohort included a substantial number of patients with refractory TED. MRI-based analyses revealed that TEP reduced both extraocular muscle size and orbital fat area, whereas IVGC conversely increased orbital fat area despite reducing extraocular muscle size. Inflamed extraocular muscles showed greater shrinkage after both TEP and IVGC therapies than non-inflamed muscles. Our findings highlight the importance of orbital fat in TED management and may contribute to optimizing treatment strategies based on MRI-detected inflammation in extraocular muscles.

## Acknowledgments

We thank Kana Hashimoto for secretarial assistance. During the preparation of this work, the authors used ChatGPT-5.5 (OpenAI; San Francisco, CA) to improve the language and readability of the text.

## Author Contributions

IY conceptualized the study. IY, MM, Yo Kishimoto, KN, and Yoshitaka Kawai designed protocols. IY, DT, and YU, TS, MM, AY, KS, EN, Yo Kishimoto, KN, and Yoshitaka Kawai obtained informed consent and collected clinical information. IY, DT, YU, TS, MA, AS, SK, DK, KO, and TH generated and verified the dataset. IY and DY supervised the study. IY drafted the manuscript. All authors discussed the results, and reviewed and commented on the drafted manuscript, and then approved the final version.

## Data Availability

Some datasets analyzed during the current study are not publicly available but are available from the corresponding author on reasonable request.

